# Supporting Reanalysis and Reuse of Clinical Trial Data: A Case Study

**DOI:** 10.1101/2025.11.06.25339683

**Authors:** Cora Burgwinkel, Han Chang Chiam, Ka Hin Tai, Jifan Wang, Mian Haider Ali, Salman Soleiman Fallah, Minoo Matbouriahi, Tobechi Obinwanne, Grigorius Papapostolou, Muhammad Riedha, Giulia Varvara, Yazid Zalai, Ulrich Mansmann, Ulrich Sax, Leonhard Held

## Abstract

**Background:** Reproducing published findings from clinical trials is a critical component of scientific transparency, yet it remains a challenging and under-practiced task. Despite increasing emphasis on reproducibility and data reuse in research policies, few real-world examples exist where independent teams have reproduced complex analyses using clinical trial data. In this case study, the aim was to independently reproduce the key findings of a high-impact clinical trial on rectal cancer treatment using shared trial data.

**Method:** We organized a multi-team datathon, where each team was provided with the same dataset and supporting material, and was tasked to reproduce the results of the CAO/ARO/AIO-04 trial, with optional additional analysis. We contacted the original investigators for data access and reuse, and consulted them to understand the study, clinically and scientifically.

**Results:** Five teams used R or Python to reproduce the statistical results, and the corresponding scripts can be found on Gitlab. All teams reproduced the analyses for primary outcome—disease-free survival (DFS). The key findings on DFS were consistently reproduced, reinforcing confidence in the trial main conclusions. Result robustness was investigated using a different analytical software or statistical models. Nevertheless, challenges were encountered when the supplementary materials were not easily identified. Minor reporting issues were noticed in the reproduced paper.

**Conclusion:** Reproduction of a major oncology clinical trial confirmed the reliability of its main conclusions. Divergences highlighted reporting gaps—such as incomplete protocols and broken links —that future trials should address. This case study demonstrates the value of systematic reproducibility checks for clinical research transparency and challenges in data sharing for reproducibility.

## 1 Introduction

Reproducing published findings from clinical trials is a critical component of scientific transparency and credibility, yet it remains a challenging and under-practiced task. Despite increasing emphasis on reproducibility and data reuse in research policies, few real-world examples exist where independent teams have successfully reproduced complex analyses using publicly shared clinical trial data [1–4]. For instance, a review of 172 data requests to the National Heart, Lung, and Blood Institute’s Biologic Specimen and Data Repository Information Coordinating Center between 2000 and 2016 found that only 2 (1%) were for the purpose of reproduction analyses [5]. Nevertheless, when such efforts are taken, reproducing a single paper can serve as a scientific check of the work and contribute to create scientific trust [1, 6]. A prominent example is the successful reproducibility study (i.e. reproduction) of the results from a cardiovascular clinical trial conducted by Gay et al. [3], which demonstrated how independent reproductions can confirm key conclusions and thereby validate the published findings. Here, reproducibility refers to the ability to consistently obtain the same results using the same data following the same analysis steps [7]. It is closely related to, but distinct from other concepts such as replicability, where similar results are obtained using different datasets, and robustness [7]. According to the International Council for Harmonisation of Technical Requirements for Pharmaceuticals for Human Use (ICH) E9 guideline, robustness is a concept which refers to the sensitivity of the overall conclusions to various limitations of the data, assumptions, and analytic approaches to data analysis, implying that the treatment effect and primary conclusions are not substantially affected when analyses are carried out under alternative assumptions or approaches [8] (see Box 1 for an overview of the definitions). These distinctions are not universally defined [9–11], and alternative interpretations have been proposed in the literature [12–14]. Reproducibility is a requirement for research credibility, as it provides the basis for assessing a study’s validity and potential for replication [15].

Efforts to improve reproducibility are increasingly supported by open science practices which promote transparency, credibility, and reusability of research through practices that make data and code openly accessible. This paradigm has gained increasing relevance in clinical research, especially with the release of the updated SPIRIT 2025 and CONSORT 2025 statements [16, 17]. These guidelines now include specific open science items to encourage more transparent planning and reporting of all relevant information from randmomized clinical trials (RCTs), such as where the trial protocol, the Statistical Analysis Plan (SAP), and the data can be accessed [16, 17]. In parallel, regulatory and industry commitments have advanced. Since 2014, biopharmaceutical companies organized through the Pharmaceutical Research and Manufacturers of America (PhRMA) and European Federation of Pharmaceutical Industries and Associations (EFPIA) have endorsed principles for responsible clinical trial data sharing, balancing accessibility with patient privacy [18]. This initiative aims to enhance the accessibility of participant-level data, clinical study reports, protocols, SAPs, lay summaries, and result publications from industry-sponsored trials [19]. Furthermore, since July 2018, manuscripts submitted to International Committee of Medical Journal Editor (ICMJE) journals that report the results of clinical trials must contain a data sharing statement, clarifying which data will be shared and with whom [20]. Together, these developments underline that transparent reporting is not only a cornerstone of open science but also an essential foundation for reproducibility, without which the validity of research findings cannot be reliably assessed [7, 9].

However, transparency alone does not guarantee that data will be reused. Despite the growing availability of shared clinical trial data, many datasets remain underutilized, often because they are neither actively requested nor reused [21, 22]. Reusing such data is essential for verifying published findings through reproduction and exploring new research questions. However, systemic barriers persist. Academic incentives still tend to favor publication volume over transparency and reproducibility [23], offering few rewards for data sharing [24]. Another challenge lies in the perceived novelty of reanalyses: often questions arise whether confirming known results is sufficient for publication, or whether reanalyses must always provide new methodological insights to be valued. Beyond those systemic barriers, there are also cultural barriers: some trialists have expressed skepticism, viewing secondary analysts as “research parasites” [25]. Such attitudes risk discouraging data sharing and reuse, sharing curated research data does not only benefit others, it can also improve the quality and credibility of one’s own work [26].

A notable example of why open science practices are essential is an RCT on chronic obstructive pulmonary disease published in 2018, which initially reported significant ben-efits from a self-management intervention. However, the article was retracted ten months later after the authors discovered a coding error during a secondary data analysis that had reversed the group assignments: what was thought to be a beneficial effect was actually harmful [27]. Over the course of the reanalysis, another minor error has been detected in imputing missing values of a secondary outcome. The corrected analysis, independently conducted by two statisticians, was later republished in JAMA [28]. This incident illustrates how easily analytical errors can go unnoticed and how rarely they are reported [29]. Had the data and code been required to be shared at publication, as recommended by open science and FAIR principles, the errors might have been detected earlier. Proper documentation, metadata, and transparent coding practices may even help prevent such mistakes, ensuring validity and reproducibility of clinical trial results.

Even with transparent documentation, data sharing, code and tool availability, true reproducibility is not automatically achieved. Reproduction still requires critical scrutiny and methodological rigor. Traditional peer review alone may not suffice to detect analytic errors or challenge questionable conclusions, as different analytical decisions can lead to different results. For instance, Ebrahim et al. [2] found that 35% of published reanalyses resulted in conclusions that differed from those of the original studies, highlighting the potential impact of methodological choices. Niven et al. [13] showed that more than half of clinical practices with a reproduction attempt demonstrated effects that were inconsistent with the original study (56%). A more recent reproduction of an oncological trial [30] demonstrated how gmethods, methods used to estimate causal effects in the presence of treatment-confounder feedback, can more appropriately adjust for treatment switching, a common issue in RCTs. The authors concluded that applying and comparing such methods can improve the validity of treatment effect estimates, especially when standard Intention-To-Treat (ITT) analyses may be biased. These examples underscore the importance of enabling multiple analytical perspectives to study the robustness of the results and to strengthen confidence in clinical trial findings.

The ability to conduct such rigorous reproductions depends on more than data availability, it also requires a well-trained workforce capable of navigating the legal, ethical, technical, and statistical complexities involved [31, 32]. Advancing this agenda depends on educating a new generation of biomedical researchers equipped with interdisciplinary expertise in data science, meta-research, and open science practices [32]. Responding to this need, SHARE and re-use Clinical Trial Data to maximise impact (SHARE-CTD) a European MSCA (Horizon-MSCA.2022-DN-01, project No. 101120360, Marie Skłodowska-Curie Actions) - funded doctoral network, provides structured professional development through formats such as interactive datathons [33]. These datathons provide hands-on, team-based learning environments in which participants collaboratively analyze shared clinical trial data to address topic-driven research questions, while gaining practical experience in reproducible workflows and the FAIRification of clinical trial datasets [32].

As part of this network, we conducted a case study to illustrate the potential of effective data reuse. The aim was to independently reproduce the key findings of a high-impact RCT on rectal cancer treatment using open-source software, such as R and Python, and shared clinical trial data. The objective was not only to replicate statistical outcomes, but also to evaluate how trial documentation, code availability, and reporting standards either facilitated or hindered our ability to reproduce the published results. To do so, we contacted the original study team for data access, organized a collaborative multi-team datathon, and engaged with the original investigators to clarify study details and reflect on the data reuse process. This work demonstrates how open science principles can be meaningfully put into practice, fostering transparency, credibility, and the broader reuse of clinical research data.

#### Key Definitions

**Reproducibility:** A result is reproducible when the same analysis steps performed on the same dataset consistently produce the same answer [7]. The analysis steps include the choice of analysis samples, variable definitions, analytical software, statistical models, and the corresponding assumptions.

**Robustness:** The concept of robustness refers to the sensitivity of the overall conclusions to various limitations of the data, assumptions, and analytical approaches to data analysis, implying that the treatment effect and primary conclusions are not substantially affected when analyses are carried out under alternative assumptions or approaches [8]. For example, a result is robust when the same dataset subjected to different analysis workflows (e.g., in R vs Python) yields similar conclusions [7].

**Reanalysis:** A reanalysis describes the act of analyzing data again, or a new analysis of data that has already been studied. It can involve repeating the original analysis to verify published results or applying alternative analytical approaches to explore the robustness of findings [1, 2].

## 2 Methods

From November 2024 to July 2025, we organized a multi-team datathon to reproduce the results of an RCT on rectal carcinoma. The datathon consisted of several online sessions and a final in-person meeting. During the online sessions, we discussed the medical background of the data and the objectives and format of the datathon, and prepared the SAP while the in-person meeting provided the opportunity to discuss programming issues, results, and conclusions.

The purpose of the datathon was to evaluate the data reuse process and to highlight the necessity of reproducibility studies. In addition to the event organizer, five teams of size two to three members were formed via random pairing, each with at least one (bio)statistician. Both the organizers (the supervisors) and the teams (the PhD students) came from the SHARE-CTD doctoral program. Each team was tasked with reproducing all the main outcomes, figures and tables reported in an original publication, with the option to extend the work with sensitivity analysis or re-analysis using the dataset.

The trial of interest was CAO/ARO/AIO-04, a German multi-center, open-label, randomized, phase 3 trial, whose main purpose was to determine whether adding oxaliplatin to then standard fluorouracil-based preoperative chemoradiotherapy and postoperative chemotherapy improved Disease-Free Survival (DFS) in patients with locally advanced rectal cancer [34]. The primary endpoint DFS was defined as the time from randomization to the first occurrence of a composite endpoint consisting of four events, namely non-radical surgery of primary tumor (R2 resection), locoregional recurrence after R0/R1 resection of primary tumor, metastatic disease or progression, and death from any cause. Randomization used block sizes of four and eight, and was stratified by study centers, clinical tumor stage, and clinical tumor nodal stage. An ITT analysis was used for the main outcome. The final results were published in 2015 [34], and we assessed its reporting quality and compared it with the CONSORT 2010 checklist [35]. Given the trial’s old publication date and hence the reduced expectation for complete open science materials, this trial provided a suitable case to assess both the facilitators and barriers of reproducibility studies.

Preparations began eight months before the on-site event (see Figure 1). We contacted the original investigators, including a medical doctor, a medical informatician, and a data scientist, of the trial in November 2024 for a collaboration agreement. The datathon was approved by the institutional review board of the University of Munich. Lectures on reproducibility practice and clinical biostatistics were provided to the teams in the previous schooling event. Anonymized data was acquired in January 2025, along with the publication in 2015 [34], a source dataset in .rda format with variables written in German, and an unofficial analysis report in .pdf format. After team formation, access to the material secured on a closed cloud storage was provided to each team. Each team was then required to complete an exercise on Git and Markdown (tools for code version control and dynamic reporting, respectively), designed to prepare members for collaborative work and good reporting practice. The original dataset was not accompanied by a data dictionary, thus a centralized data dictionary was prepared by the event organizer and provided to the five teams in April, in particular translating some variables from German to English. After preliminary exploration of the dataset, a Q&A session was held online in mid-May for the team members to discuss their findings and ask questions. The first version of the SAP was submitted by each team, assessed by the organizer, and presented in an internal web conference in June. The five teams were free to decide how they approach the tasks, e.g. coding languages and selection of variables. To ensure data protection, the work of the five teams was anonymized, re-rendered in a new Git workspace, and the dataset and scripts were adapted for reproduction purposes only. Additional analyses, using variables which have been judged by the group of higher risk of patient re-identification, were removed from our manuscript.

**Figure 1:**
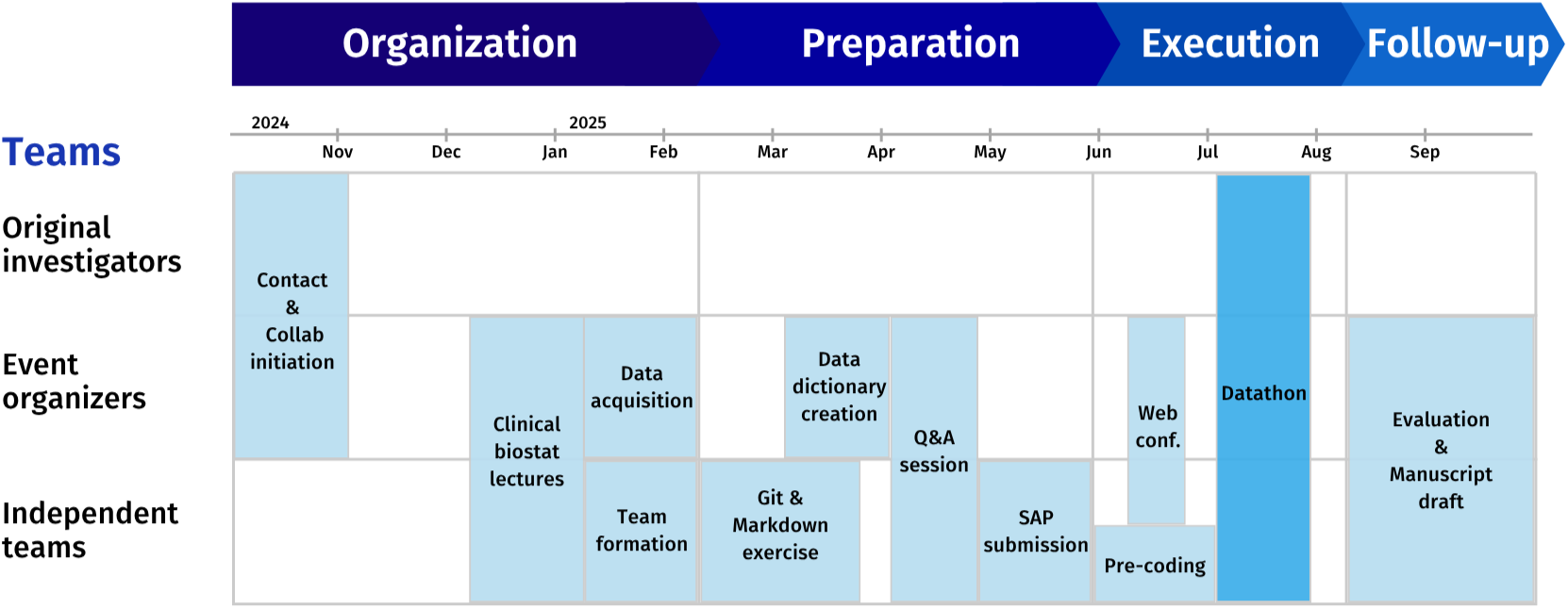
Timeline of the Datathon

The on-site datathon lasted for five days in July 2025 in Göttingen, Germany. Apart from numerous sessions of team coding, several lectures were also given to the teams by our principal investigators (UM, LH) on topics like survival analysis, methodological considerations, and rationale for approach selection. A clinician and a study coordinator from the original investigators were invited to lecture the teams about the clinical detail of the trial and to explain their analytical considerations in the original study. At the climax of the datathon, each team presented their reproduction results and other teams were instructed to raise questions on methodology and analytical decisions as well as interpretation. The original investigating clinician also engaged in the discussion and provided further insights to the study dataset. The expected outputs were an analysis Markdown report, a Git workspace with the coding script, the material for reproduction, and the presentation of results. A prize was awarded to the best-performing team, assessed by their script, reports, and presentation. An evaluation session followed involving all teams and the coordinating organizer, where we reflected on our experience throughout the whole data reuse process and during the conduct of the reproducibility study, highlighting both positive takeaways and challenges.

## 3 Results

The primary goal of all teams was to reproduce the results of the primary outcome and the results presented in the tables and figures in the publication, including baseline characteristics, incidence rates of efficacy endpoints, death cause profile, the CONSORT diagram, Kaplan-Meier curves of DFS and Overall Survival (OS), cumulative incidence curves of locoregional recurrences and distant recurrences, and the forest plot of subgroup analyses on DFS (see Table 1). The sample size calculation reproduction was attempted together with one of the principal investigators (LH) during the biostatistics session of the datathon. Each team proposed distinct additional analyses (see Table 2) in their SAPs before the datathon. Four teams used R for analysis as in the original study, and one team (Team 5) used Python. The primary hypothesis testing was based on the coin package in R, which is not available in Python. Different software can yield different results due to model availability and/or programming differences, which will be further discussed in section 3.2. The non-statistical R package versions, visualization package/functions, and parameters used in specific functions varied across teams. Our statistical analysis plans and reproduction scripts are available on GitLab and Zenodo.

**Table 1:**
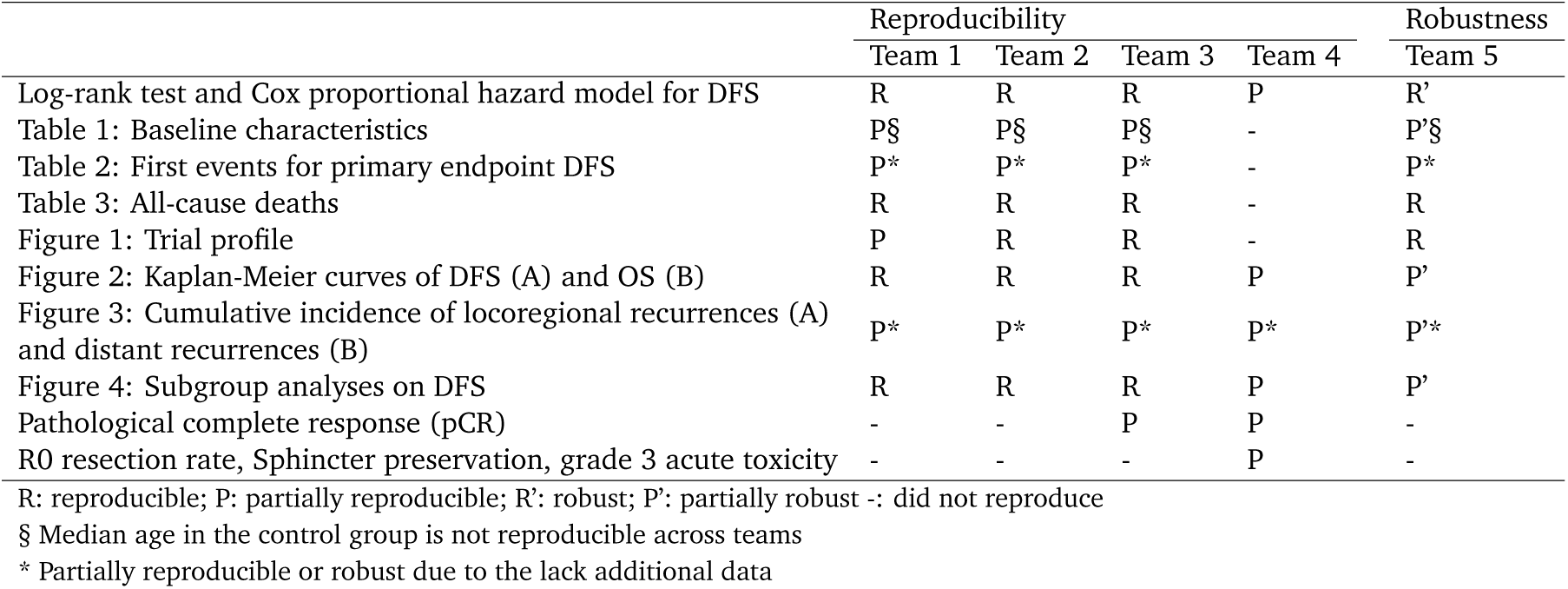
Reproducibility assessment from multiple teams.

**Table 2:** Robustness check and exploratory analysis from multiple teams.

|  | Team 1 | Team 2 | Team 3 | Team 4 | Team 5 |
| --- | --- | --- | --- | --- | --- |
| Additional analyses |  |  |  |  |  |
| Robustness: statistical models | N | Y | Y | N | Y |
| Robustness: missing values | N | N | N | Y | Y |
| Robustness: software (other than r) | N | N | N | N | Y |
| Refinement of original results | Y | Y | Y | N | Y |
| Exploratory analyses with potential clinical impact | N | Y | Y | Y | Y |

Y: Implemented; N: Not implemented

### 3.1 Reproduction

Considering how many same results were obtained using the same analysis method, our reproduction was successful. Most teams were able to reproduce all figures and tables with a handful of minor numerical differences, except for median age in the control group and the cumulative incidence of locoregional recurrence (see Table 1). The teams discovered later that an additional 19 data records on time to locoregional recurrence were added in the original analysis script and were not part of the data shared with us. One team could not reproduce all the sample sizes along the intervention procedures due to the wrong choice of procedure variables. One team had an issue including the last year data in the Kaplan-Meier curves of DFS and OS; another team could not fully reproduce the number of participants at risk for cumulative incidence or locoregional and distant recurrences, possibly due to the handling of ties. One team yielded discrepant results in some subgroups due to the handling of the clustering variable. Even though the reproduced trial was published 10 years ago, and the teams did not see the original SAP or original data dictionary during the reproduction, most results were reproduced identically by at least two teams. Therefore, we consider the reproduction a success.

The teams identified some discrepancies between the data and the reported categories of participant characteristics in the publication during the reproducibility assessment. The reported categorization of some subgroups is different from the one the teams used in the reproduction. For example, using the reported age groups, we obtained different hazard ratios in the subgroup analysis. After shifting one year between two categories, we were able to reproduce the reported results. A similar issue is observed for location from anal verge and pathological T stage (ypT). These misreportings hindered our reproduction; moreover, it may mislead the clinical interpretation of the results.

Among all the secondary outcomes of the trial, two teams calculated the pathological complete response (pCR) rates with a logistic regression, and one team attempted to reproduce the results of locoregional (R0) resection, sphincter-sparing resection, and postoperative grade 3 or higher acute toxicity. The pCR rates are reproducible, while the odds ratio and its confidence interval are not. The teams encountered the convergence issue when applying the mixed-effects logistic model (i.e., the counterpart of the analysis for the primary outcome), so they used the fixed-effect logistic model instead. Limited information on secondary outcome analysis methods is found in the article, so we excluded secondary outcome testing and modeling from the reproducibility assessment and will discuss them further in the data reuse section. Our analysis yielded discrepant rates of R0 resection and sphincter-sparing resection, but the same rate of postoperative grade 3 or higher acute toxicity.

#### 3.1.1 Trial Documentation

The main trial document used by the five teams for reproduction is the trial primary publication [34] and an unofficial analysis report provided by the investigators. The unofficial analysis report contains some, not all, analytical decisions that were not reported in the paper, for example, the usage of a collapsed clustering variable in the log-rank test because of small clusters. Additional analytical details, such as the seed number used for the log-rank test, are unavailable. A few documents were not available during the process of reproduction, including the original data dictionaries, the trial protocol in English, and the original SAP. A brief data dictionary was generated by the datathon organizer and provided to us, but the information is limited, especially on the processed variables. The trial protocol was linked in the publication, but the link was invalid when the teams tried to access it. The trial protocol and its English Synopsis were found available on a website through a search during the datathon event. The full protocol contains the original SAP.

#### 3.1.2 Code Availability

The code was shared through the aforementioned invalid link in the publication. Investigators also shared the analysis code with the datathon organizer. Due to the educational purpose of this datathon, the five teams did not receive the code during the reproducibility assessment.

#### 3.1.3 Reporting Standards

Like most reproducibility studies, our reproduction mainly relied on the reporting from the publication of the original study; thus, we evaluated the publication with the CONSORT 2010 checklist (Supplementary Material Table S1). Out of 37 items, we were able to identify the clear reporting of 24 items and partial reporting on 6 items. Three items are irrelevant to the study, while two items are unable to assess because of no access to the protocol by the time of datathon. Another two items were reported yet incorrect, or no information could be obtained based on the reporting.

Among all the items, two are particularly relevant to our reproduction—the reporting of how the sample size was determined (item 7a) and the location where the full trial protocol can be accessed (item 24). The sample size calculation was not reproducible solely based on the publication. During the datathon event, we found that the sample size calculation is clear in the protocol (i.e., Prüfplan in German) and that the reporting on the sample size determination is partially inaccurate in the publication. The protocol was linked in the publication, but the link was broken when we attempted to access it.

Other reproducibility-related reporting issues are not easy to identify based on the CONSORT checklist. The hypothesis testing on the primary outcome was based on a stratified log-rank test, as proposed in the protocol. Some levels of the stratified factor were collapsed in practice due to the small number of participants in some strata. On the other hand, the pre-collapsed stratified factor was used in the mixed-effect models for the primary outcome. Another specific issue is the unknown seed number used in the log-rank test, required by the reported package (i.e., coin). Although all teams reached the same test conclusion, the exact p-value was not reproducible. Computational environment issues also require attention. One team tried to downgrade R to use the packages in the reported version, and was stopped by the issues of CPU compatibility and limited R markdown functionality in the old version. This level of detail can be easily overlooked in the reporting, even though the statistical methods were generally clear in the publication.

### 3.2 Robustness

In line with ICH E9 guideline, we assessed robustness by exploring sensitivity of the trial conclusions to alternative analytical approaches. This included software robustness, where conclusions were compared across implementations in R and Python (Table 2). The first type of assessment focused on the statistical assumptions of the implemented model, e.g. mixed-effects Cox models that depend on exchangeability within the stratification factor; different stratification factors were explored by two teams. Different specifications for the Cox model were fitted for DFS and OS, including a fixed-effects model without stratification, a fixed-effects model with center entered as a factor, a mixed-effects model with a random intercept for center, and a mixed-effects model stratified by clinical tumor stage. Both teams obtained very similar hazard ratio estimates and overlapping confidence intervals across models. For an additional secondary endpoint (pathological complete response), the generalized linear mixed model analogous to the primary analysis did not converge. An unplanned stratified Cochran–Mantel–Haenszel test was reported in the trial publication, and one team also conducted simple Chi-square and logistic regression analyses, which yielded similar results. Furthermore, the possible impact of missing magnetic resonance imaging assessments in clinical tumor stage and clinical nodal stage was explored by one team. The full analysis set was also evaluated after multiple imputation of MRI based clinical tumor stage and clinical nodal stage, with comparison to original analyses which simply utilized the worst outcome from the actually available assessment methods.

Robust results are those that do not depend on the specificities of the programming language chosen to perform the analysis [7]. Software robustness was checked through implementation in R by four teams and in Python by one team. Besides the difference in the programming language, stratified Cox regression is used in place of the mixed-effects Cox model as the package of choice does not support mixed-effect Cox model. Modules in Python also do not support the stratified exact log-rank test for hypothesis testing of the primary endpoint, so the team proceeded with the stratified log-rank test. Most results are in line with the conclusion from the paper. Taken together, these exercises show where conclusions remain stable and where model choices, assumptions about missing data, or software environments influence the results.

### 3.3 Reanalysis

Distinct analyses by five independent teams in this case study demonstrated added value by reanalyzing the data, as summarized in Table 2. Refinement of the published results in Rödel et al. [34] was made with a newer R package (i.e., survminer). For instance, confidence bands were added to Kaplan Meier curves and to cumulative incidence functions. The original investigators published a forest plot presenting hazard ratios for DFS in various subgroups with 95% confidence intervals. Beyond this, one team further assessed the evidence for heterogeneity of treatment effects in subgroups using an interaction test. These small adjustments made uncertainty explicit, extended the clinical information of DFS and OS beyond the original report, and followed current standard on the reporting of subgroup analyses in clinical trials [36].

Additional exploratory analyses provided insights in clinical context. For example, time trend patterns in baseline characteristics across the recruitment period were assessed by one team to detect potential imbalances that might contribute to variation in the treatment effect over time. Another team summarized counts of deaths by surgical method and by center to screen for heterogeneity that could merit further investigation. These additional explorations and analyses illustrate how shared data support questions beyond strict reproducibility.

## 4 Discussion

This case study demonstrates that reproducing the findings of an RCT using shared data is not only feasible, but also provides meaningful scientific value. Each reproduction effort contributes to strengthening the reliability of the scientific record, either by validating the original findings or, in some cases, by revealing discrepancies that warrant further investigation. Our study benefited greatly from the substantial time commitment of the teams involved, whose dedication enabled a thorough and detailed reproduction of the trial, and by the access to a clinician and a study coordinator involved in the original study. The multi-team approach further enhanced the process by fostering discussion, particularly around methodological rationale and interpretation of results, which in turn enriched the analytical process and deepened our understanding of the original work.

This work goes beyond a reproducibility exercise by offering a reflection on how clinical trial data owners, sponsors, and publishers can better support reproducibility and data reuse in practice. Notably, the original study team was under no obligation to share the dataset or respond to our request, yet their willingness to engage was essential to the success of this project. Moreover, the clinician and the study coordinator also provided a lecture on the essential clinical context to us to help our reproduction. Such openness should be encouraged and incentivized to show the scientific community the full potential of shared clinical trial data.

Our findings support previous reviews, which noted that many trial groups face challenges when sharing research data [1, 24]. Building on these challenges, we identified broader, recurring issues in current data-sharing practices. The lack of transparent reporting is a widely discussed yet lingering issue. The SPIRIT 2025 and CONSORT 2025 place greater emphasis on reporting practices that support open science compared to their previous versions, requiring study protocols and publications to reference the locations of trial protocol, SAP, de-identified participant data, data dictionary, and statistical code [16, 17]. Because of the time of the original study, we compared the study publication with CONSORT 2010 [35] instead of CONSORT 2025 [17]. Although, as shown in our comparison, the study publication adheres well enough to the CONSORT 2010, this alone doesn’t ensure reproducibility. For example, the link to the protocol, SAP, and analysis code was broken, preventing full verification of the reported analyses. The storage of trial datasets on private initiatives versus in sustainable and long-term repositories can raise concerns about the persistence and accessibility of these resources over time. Ensuring reproducibility requires accessible datasets and scripts, and the provision of the complete computational environment, including software versions, dependencies, and configurations. Ongoing studies aim to evaluate the effectiveness of open science interventions from all research stakeholders [37] and to establish a reproducibility checklist to guide the open-science reporting beyond the current guidelines [38]. As more researchers report trial registry numbers in the publications [39, 40], trial registries may become an additional resource for reproduction. Unfortunately, trial registries often provide only limited supplementary information, which is insufficient when the published paper lacks methodological clarity.

Addressing these issues requires structured and transparent data sharing practices that go beyond simply providing datasets. The FAIR principles [41] should be adopted to enhance data sharing and long-term accessibility. Among the processes involved in data FAIR-ification [42], the most crucial elements for supporting reproducibility are comprehensive metadata describing both the study and the dataset, along with a standalone data dictionary detailing the variables. Supplementary materials should also be made available whenever possible, such as study protocols, SAPs, analytic code, and original case report forms, so that methodology rationale, variable definitions, and analytic decisions can be fully understood and verified. The computing environment can be provided through a virtual machine, a container image (of the operating system), or a package/environment management system, along with the analysis script [43]. If not possible, the software version, dependencies, and configurations should be documented as a minimum. Such documentation, when provided with the dataset, enhances reproducibility and creates opportunities for secondary analyses and broader data reuse.

Secondary use of clinical data can benefit from advancements in both methodology and clinical knowledge. Many trials originally reported complete case analysis, while imputations can now possibly be informed by improved statistical techniques with a better understanding of disease mechanisms and prognostic factors that influence missingness. Secondary analyses of subgroups and additional endpoints may also provide insights that were not accessible at the time of the original publication.

Artificial intelligence (AI) may also enable automation of reproducing published clinical trial results. With well documented datasets and clear SAPs, AI systems could re-execute analyses and validate reported findings. This capability would be extremely valuable in large-scale reproduction efforts, such as the example from economics and political science [44]. Tools such as CONSORT-NLP [45] and the more recent RAPID framework [46], which automatically generate trial reporting checklists, show that AI can support reproducibility efforts by improving reporting consistency and validation of protocol adherence. The recently developed Reproducibility Copilot illustrates how AI can automate computational reproduction: it analyzes manuscripts, code, and supplementary materials to generate structured Jupyter Notebooks, reducing reproduction time from more than 30 hours to about 1 hour while systematically detecting barriers to reproducibility [47]. Although there are not yet documented cases in clinical trials where AI fully reproduces published findings, these developments show the technical feasibility and potential of AI to support validation, robustness checks, and exploratory analyses once original trial data and analytic materials are accessible.

The question of how to better use reproducibility studies remains unanswered. The traditional publications in academic journals are not suitable for connecting pieces of information together for reasons like pay wall, page limit, multiple publications from one trial data, etc. In order to track which trials were reproduced, trial registries have the potential to serve as central information hubs of clinical trials, linking the reproductions to the original trials, as reporting the trial registration number is often required at publication. Logistic issues, such as who can and will add this information, are unclear. Trial registries also suffer from the issue of poor result reporting practice [48], which limits the extent to which this potential is likely to be realized in the near future. Complementing the track of reproductions, transparent reporting in reproducibility and reanalysis studies is equally important. To our knowledge, there is no clear guide on how to report those types of studies. In our case study, we didn’t provide a numeric side-to-side comparison with the original study because we have five independent reproductions on the same trial based on the educational purpose of our datathon, but all the scripts are available on the Gitlab and Zenodo for transparency. Reproducibility studies usually present results from the reproduction along with the original paper in the format of tables or forest plots [3, 6, 49], which is straightforward for readers. The reporting should follow the same standards as in the original study, with additions on the process of identifying data and supplementary materials.

Reproducing a single paper is already a meaningful and relevant contribution to science. Each successfully reproduced study adds to the collective body of evidence that can be trusted, helping to strengthen the foundation upon which future research, clinical guidelines, and policy decisions are built. Beyond verifying specific findings, reproduction fosters a culture of accountability and rigor, sending a clear signal that published work is open to scrutiny and confirmation. This, in turn, can encourage higher reporting standards, better documentation, and more transparent data sharing from the outset of research projects.

The benefit extends beyond the original study: each reproduction effort contributes to methodological learning, identifies potential pitfalls, and demonstrates reproducible workflows that others can follow. Ultimately, the cumulative effect of multiple reproducible studies is greater scientific robustness, enhanced public trust, and improved decision-making in healthcare and beyond.

## Data Availability

The data from the CAO/ARO/AIO-04 trial are available in the R package TH.data and can be assessed as detailed in the R code which we uploaded to Gitlab and Zenodo.

## List of abbreviations

CONSORT: Consolidated Standards of Reporting Trials
DFS: Disease-Free Survival
ICH: International Council for Harmonisation of Technical Requirements for Pharmaceuticals for Human Use
ITT: Intention-To-Treat
OS: Overall Survival
RCT: Randomized Clinical Trial
SAP: Statistical Analysis Plan
SHARE-CTD: SHARE and re-use Clinical Trial Data to maximise impact
SPIRIT: Standard Protocol Items: Recommendations for Interventional Trials

## Declarations

### Ethics approval

For our project, we submitted a waiver approval to the ethics committee at the University of Munich. The committee evaluated the provided information. The ethics committee approved the study (Project No.: 25-0750-KB) and raised no objections to its conduct.

### Consent for publication

Not applicable

### Availability of data, code and materials

The data from the CAO/ARO/AIO-04 trial are available in the R package TH.data and can be assessed as detailed in the R code. The R code for the reproduction presented in this work can be accessed in the Gitlab and Zenodo. This repository includes all the scripts used for this manuscript.

## CRediT authorship contribution statement

**Conceptualization**: CB, HC, KH, JW, LH, UM, US

**Methodology**: CB, HC, MH, GP, GV, KH, MM, MR, TO, UM, JW, LH

**Software**: CB, HC, MH, GP, GV, KH, TO, JW, YZ, LH

**Validation**: CB, HC, MH, GV, KH, MM, MR, TO, YZ

**Formal analysis**: CB, HC, MH, GV, KH, TO, JW, YZ, UM, LH

**Investigation**: CB, HC, KH, JW, MH, SF, TO, MM, GP, MR, GV, YZ, LH, UM, US

**Resources**: LH

**Data curation**: US, SF

**Writing - original draft**: CB, HC, KH, JW, LH, UM, US

**Writing- review & editing**: CB, HC, KH, JW, MH, SF, TO, MM, GP, MR, GV, YZ, LH, UM, US

**Visualization**: CB, HC, MH, GP, GV, KH, MM, MR, TO, JW

**Supervision**: LH, UM, US

**Project administration**: LH, UM, US

**Funding acquisition**: LH, UM, US

## Acknowledgements

The authors would like to thank the investigators and the study team of CAO/ARO/AIO-04 for providing access to the clinical trial dataset and their support in clarifying study details. We specifically thank Torsten Hothorn for preparing and curating the dataset, and Torsten Liersch and Johanna Kreutzer for sharing essential clinical context through their lectures.

---

**Supplementary Table S1:**
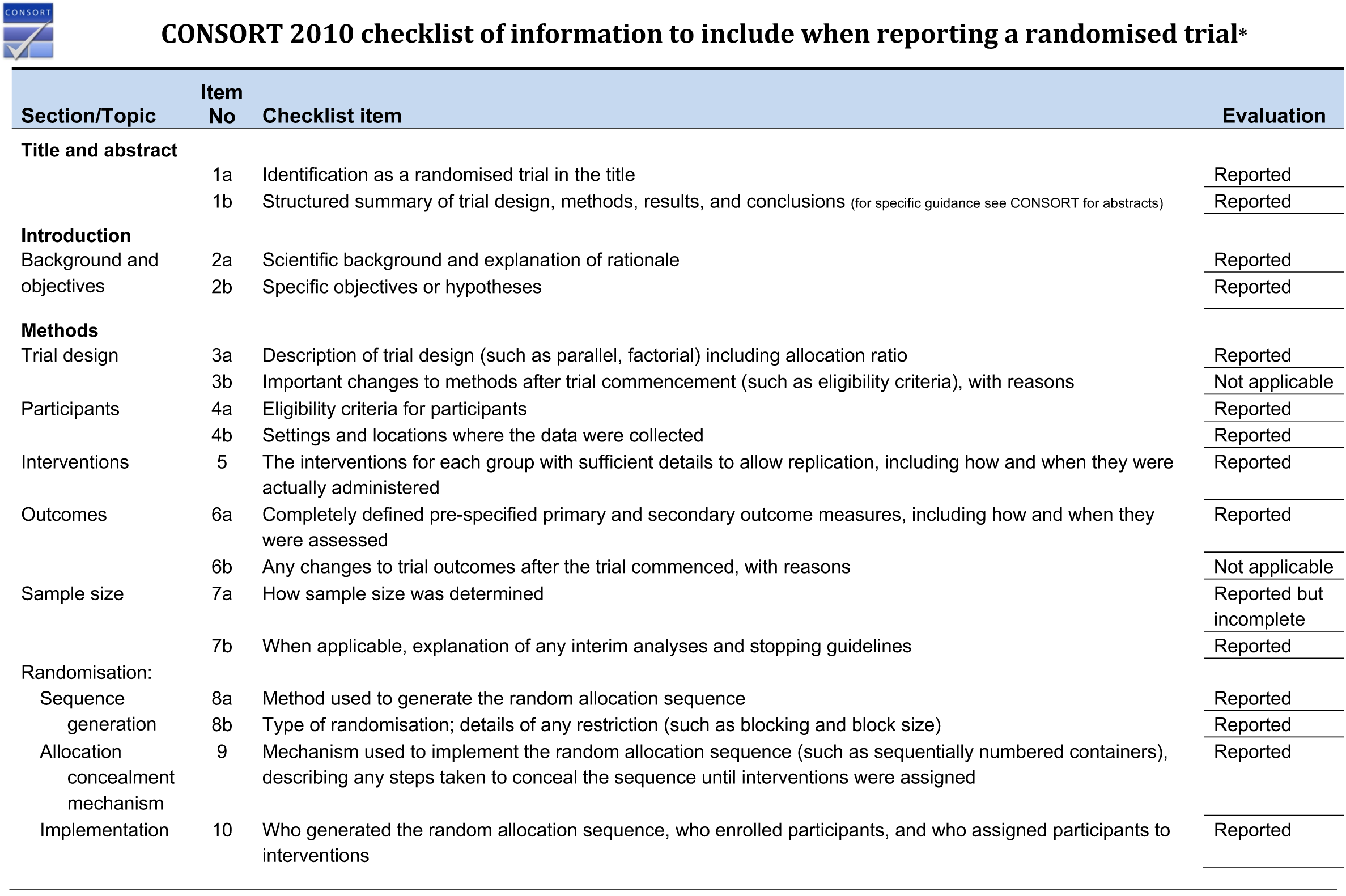
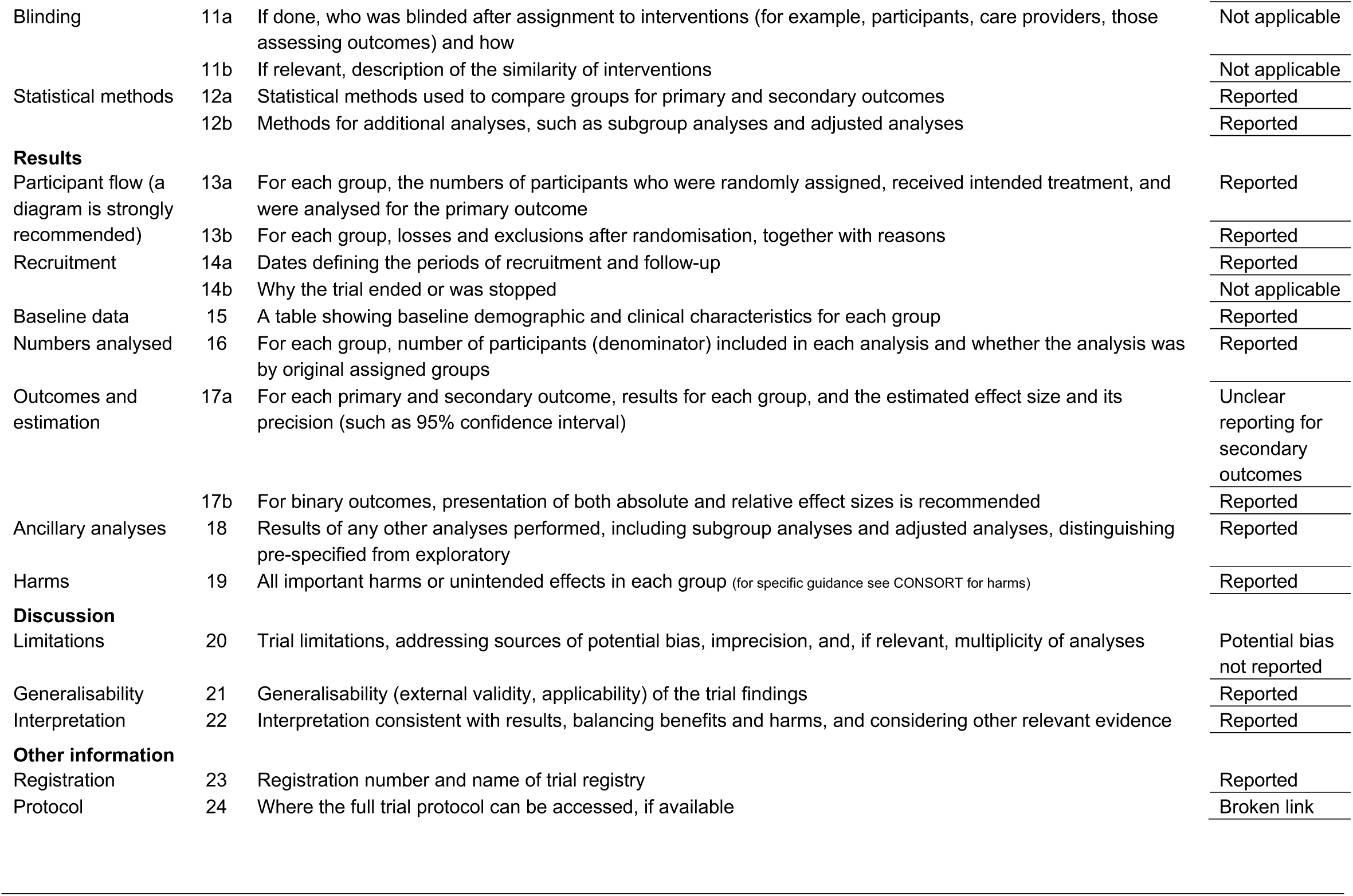
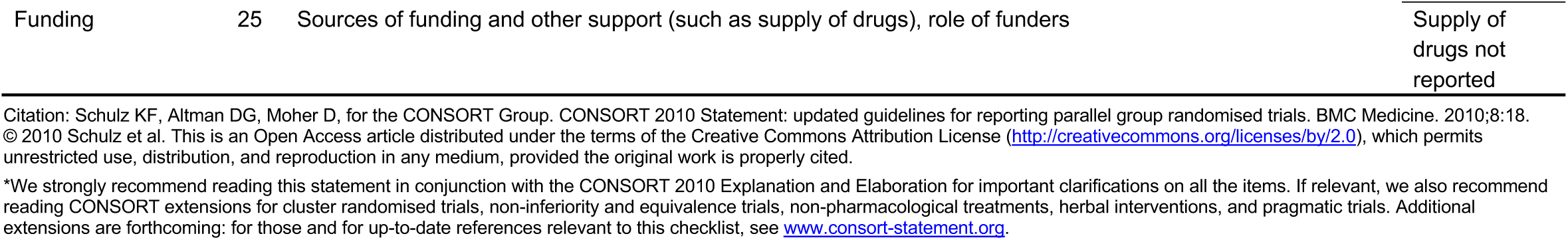
Evaluation on the reproduced paper according to CONSORT 2010 checklist.

